# Maternal Vitamin B12 Deficiency and perinatal and Child Neurodevelopmental Outcomes: A Systematic Review and Meta-Analysis

**DOI:** 10.64898/2026.10.07.26364974

**Authors:** Eduarda Rodrigues Belfort de Carvalho, Hércules Kanaan Pereira Sousa, Thiago de Sousa Siqueira, Ricardo Rezende de Queiroz, Juliana Borges Tavares da Silva, Ana Júlia Hortegal Cavalcante, Paloma Layonara Vieira Viana, Maria Eduarda Araújo Magalhães, Gabriel Segrini Fidencio, Ana Raquel Batista de Carvalho

**Affiliations:** Centro Universitário UNINOVAFAPI, Brazil

**Keywords:** Vitamin B12, cobalamin, pregnancy, preterm birth, birth weight, small for gestational age, neurodevelopment, meta-analysis

## Abstract

**Background:** Vitamin B12 is essential for one-carbon metabolism, deoxyribonucleic acid (DNA) synthesis, and myelination, and inadequate maternal status during pregnancy may influence fetal growth and child neurodevelopment.

**Objective:** To evaluate the association between maternal vitamin B12 status and perinatal and child neurodevelopmental outcomes.

**Methods:** We conducted a systematic review and meta-analysis of observational studies and randomized clinical trials. The main outcomes were preterm birth, low birth weight, small for gestational age (SGA), and cognitive/global development. For binary outcomes, the natural logarithm of the relative risk (logRR) was the primary analytical effect measure; for neurodevelopment, Hedges g was used. Random-effects models were fitted, between-study variance was estimated by restricted maximum likelihood (REML), and inference used Knapp-Hartung (KH) adjustment.

**Results:** For preterm birth, nine estimates yielded a pooled logRR of 0.190 (95% confidence interval [95% CI] KH: –0.046 to 0.427; p = 0.101; I2 = 19.2%). For low birth weight, nine estimates yielded a pooled logRR of 0.136 (95% CI KH: –0.013 to 0.286; p = 0.069; I2 = 4.8%). For SGA, six estimates yielded a pooled logRR of 0.047 (95% CI KH: –0.083 to 0.177; p = 0.397; I2 = 0%). Cognitive/global development, assessed in five independent studies, showed a pooled Hedges g of 0.266 (95% CI KH: 0.034 to 0.499; p = 0.034; I2 = 55.7%).

**Conclusions:** logRR estimates for preterm birth and low birth weight were directionally compatible with higher risk among mothers with lower vitamin B12 availability, but their 95% CIs included the null. No consistent association was observed for SGA. Higher maternal vitamin B12 availability or supplementation was associated with a small favorable effect on cognitive/global development, with moderate heterogeneity.

## 1 INTRODUCTION

Vitamin B12, or cobalamin, is an essential micronutrient involved in several metabolic processes, including DNA synthesis, cell division, one-carbon metabolism, methionine formation, and maintenance of myelin. During pregnancy, these functions are particularly important because of intense cellular proliferation, placental and fetal growth, and rapid maturation of the central nervous system. Inadequate maternal vitamin B12 availability may therefore affect both fetal development and neurological functions that become evident later in childhood. Previous studies have reported inconsistent associations between maternal B12 status and perinatal outcomes, reinforcing the need for a quantitative synthesis that accounts for differences across populations, study designs, and effect measures (Green *et al*., 2017; Pepper and Black, 2011).

Maternal vitamin B12 deficiency during pregnancy may result from insufficient dietary intake, low availability of animal-source foods, gastrointestinal malabsorption, metabolic changes, or increased physiological requirements. Populations following vegetarian or vegan dietary patterns are at particular risk when supplementation is not adequate, although deficiency may also occur among omnivorous women. In addition to diet, socioeconomic conditions, supplement use, preconception nutritional status, and interactions with other micronutrients involved in one-carbon metabolism may modify maternal concentrations throughout pregnancy (Allen, 2009; Green *et al*., 2017).

From a pathophysiological perspective, vitamin B12 participates in the conversion of homocysteine to methionine and in the regeneration of tetrahydrofolate, steps related to nucleotide synthesis and cellular methylation. Disturbances in these pathways may influence placental growth, angiogenesis, cell proliferation, and epigenetic regulation. In parallel, B12 is required for maintenance of myelin integrity and neuronal metabolism. These mechanisms provide biological plausibility for investigating its relationship not only with immediate obstetric outcomes but also with cognitive, motor, and language development throughout childhood (Green *et al*., 2017; Pepper and Black, 2011).

Among the most commonly studied perinatal outcomes are preterm birth, low birth weight, and small-for-gestational-age birth. These events share some determinants but represent distinct phenomena. Low birth weight may result from either shortened gestation or insufficient fetal growth, whereas classification as small for gestational age considers birth weight in relation to gestational age. Evaluating these outcomes separately is therefore important to distinguish possible effects of maternal vitamin B12 deficiency on gestational duration and intrauterine growth (Rogne *et al*., 2017).

The available evidence, however, is not uniform. Observational studies conducted in different populations have reported associations of varying magnitude between maternal B12 concentrations and preterm birth or birth weight, whereas others have not identified consistent relationships. Similarly, preconceptional or gestational supplementation trials have shown heterogeneous results, reflecting differences in dose, timing of initiation, baseline B12 status, maternal diet, and clinical characteristics. This variability complicates the interpretation of isolated findings and supports the use of models capable of incorporating clinical and methodological heterogeneity (Bonifacio *et al*., 2025; Rogne *et al*., 2017).

Neurodevelopment represents an additional dimension of this relationship. Fetal exposure to vitamin B12 deficiency occurs during a period of rapid structural and functional organization of the brain, and its effects may not be fully captured by anthropometric measures at birth. Studies following children after pregnancy have used instruments such as global developmental scales, cognitive measures, motor assessments, and language tests. This methodological diversity makes direct comparisons difficult, but also expands the possibility of detecting effects that emerge months or years after gestational exposure (Bhate *et al*., 2012; Bonifacio *et al*., 2025; Lai *et al*., 2019).

Another challenge concerns the heterogeneity of definitions of vitamin B12 deficiency. Different studies used serum or plasma concentrations, distinct cut-off points, and measurements taken at different moments of pregnancy. In addition, some studies evaluated observational exposure, whereas others investigated maternal supplementation. These approaches address related but not identical questions, requiring careful harmonization of effect measures and explicit consideration of between-study heterogeneity (Bonifacio *et al*., 2025; Rogne *et al*., 2017).

In this context, this systematic review and meta-analysis aimed to evaluate the association between maternal vitamin B12 status and perinatal and child neurodevelopmental outcomes. The main analyses were structured for preterm birth, low birth weight, SGA, and cognitive/global development, using random-effects models, REML estimation of heterogeneity, and KH inference. For the three binary outcomes, logRR was defined as the primary analytical effect measure. Secondary outcomes related to neonatal anthropometry and motor and language development were also considered when available.

## 2 METHODS

### 2.1 Study design and research question

We conducted a systematic review with meta-analysis to evaluate the association between maternal vitamin B12 status and perinatal and child neurodevelopmental outcomes. The review was structured according to the Preferred Reporting Items for Systematic Reviews and Meta-Analyses (PRISMA) recommendations. The research question was organized according to population, exposure/intervention, comparator, and outcomes. Eligible populations included pregnant women evaluated for vitamin B12 status or receiving vitamin B12 supplementation, as well as their conceptuses and offspring followed during the neonatal period or childhood. The analytical protocol included observational studies and randomized clinical trials (Page *et al*., 2021).

The main outcomes were preterm birth, low birth weight, SGA, and cognitive or global neurodevelopment. Head circumference, motor development, language, and other neonatal parameters were considered secondary outcomes when reported and remained in narrative or exploratory synthesis when the number or homogeneity of estimates was insufficient for quantitative pooling.

### 2.2 Search strategy and study identification

The bibliographic search included PubMed/MEDLINE (Medical Literature Analysis and Retrieval System Online), Scopus, Latin American and Caribbean Health Sciences Literature (LILACS), and Google Scholar. Controlled descriptors and free-text terms related to vitamin B12, cobalamin, pregnancy, maternal deficiency, supplementation, preterm birth, birth weight, fetal growth, SGA, and child neurodevelopment were used. Terms were combined using Boolean operators according to the syntax of each database.

Targeted searches were also conducted to identify primary studies with quantitative results compatible with the predefined outcomes. References of eligible studies and publications with individual participant data syntheses were examined to locate potentially relevant primary cohorts. When an individual participant data meta-analysis provided study-specific, standardized estimates for each original cohort in published supplementary material, those estimates were used as the quantitative source for the respective cohort, without counting the secondary meta-analysis as an additional primary study (Rogne *et al*., 2017).

Study selection was performed in two stages: first by screening titles and abstracts and then by full-text assessment of potentially eligible reports. Data extraction and methodological quality assessment were performed by two reviewers. Disagreements were resolved by consensus.

### 2.3 Eligibility criteria

We included studies evaluating maternal vitamin B12 concentrations during the preconceptional or gestational period, maternal deficiency defined by specific laboratory criteria, or vitamin B12 supplementation administered before or during pregnancy. Prospective and retrospective observational studies, birth cohorts, and randomized clinical trials were eligible.

For quantitative inclusion, studies had to provide sufficient data to obtain an effect measure and its variance, either through a relative risk and corresponding confidence interval, event counts and denominators, means and standard deviations, standardized coefficients, or other statistics amenable to transformation. For binary outcomes, all eligible estimates were converted to logRR before pooling.

We excluded publications without an interpretable comparator, studies without maternal or gestational vitamin B12 assessment when the purpose was to infer maternal effects, case reports, reviews, editorials, and studies without usable quantitative information. When a publication reported relevant results in a format incompatible with the main meta-analysis, data were retained for narrative synthesis.

### 2.4 Data extraction and harmonization

For each study, we recorded authorship, year, country, design, sample size, maternal characteristics, timing of vitamin B12 assessment, deficiency criterion, supplementation when applicable, gestational age, newborn characteristics, developmental assessment instrument, and quantitative results.

Binary outcomes – preterm birth, low birth weight, and SGA – were harmonized directly on the logRR scale, defined as the natural logarithm of the relative risk. This was the scale used for estimation, weighting, presentation of individual effects, and calculation of pooled effects. When a study provided relative risk and 95% CI, both interval limits were transformed by the natural logarithm and the standard error was derived on the logRR scale. When events and totals were available for each group, the relative risk was first calculated and then logarithmically transformed. Exponentiation of logRR was reserved only for complementary clinical interpretation and did not constitute the primary effect measure of the meta-analysis.

The direction of comparisons was standardized so that logRR values above 0 represented a higher frequency of the adverse outcome associated with deficiency or lower maternal vitamin B12 availability, whereas 0 represented no association. When a publication used the group with higher B12 as the reference, the contrast was inverted before transformation to maintain the same analytical orientation.

Cognitive/global development was harmonized as a standardized mean difference with Hedges correction, expressed as Hedges g. When means, standard deviations, and sample sizes were available, the standardized difference was calculated directly. Estimates originally reported in standard deviation units or compatible coefficients were converted to the same scale when mathematically justified. Positive values were oriented to indicate better neurocognitive performance associated with higher maternal vitamin B12 availability or supplementation.

### 2.5 Statistical synthesis

Meta-analyses were conducted separately for each outcome. Random-effects models were used, considering expected differences across populations, deficiency criteria, exposure timing, supplementation, assessment instruments, and follow-up duration.

Between-study variance (tau2) was estimated by REML. Inference for the pooled effect used KH adjustment, which incorporates uncertainty related to heterogeneity estimation and produces more conservative confidence intervals in datasets with a limited number of studies. Pooled effects, 95% CI, and p values were reported.

Heterogeneity was assessed using Cochran Q, I2, and tau2. For the three binary outcomes, individual effects, intervals, and pooled estimates were maintained in logRR in all main quantitative presentations. Exponential transformation was used only as a complementary interpretive resource. For cognition, effects were presented as Hedges g.

When a study provided a reconstructable crude statistic but its published interpretation depended on an adjusted model not described sufficiently for conversion, that estimate was not automatically incorporated into the main analysis. In these cases, the result was reserved for sensitivity analysis or narrative synthesis. Forest plots for the four main outcomes were placed in the appendices, while the manuscript body focused on pooled estimates, heterogeneity measures, and the principal quantitative interpretation.

## 3 RESULTS

### 3.1 Study selection

The database searches identified 2,107 records. After removal of 681 duplicate records, 1,426 records underwent title and abstract screening, of which 1,198 were excluded. Full-text reports were sought for 228 records; seven reports could not be retrieved, leaving 221 reports for full-text eligibility assessment. Of these, 208 reports were excluded: population outside the eligibility criteria (n = 41), ineligible or non-isolable vitamin B12 exposure/intervention (n = 43), absence of an eligible perinatal or neurodevelopmental outcome (n = 47), ineligible study design (n = 25), insufficient quantitative data for extraction or meta-analysis (n = 22), secondary publication or overlapping study population (n = 12), and review, editorial, letter, protocol, or other non-original publication (n = 18). Thirteen studies were included in the review. The complete PRISMA 2020 study-selection flow diagram is presented in Figure E1 (Appendix E).

### 3.2 Composition of the quantitative synthesis

The quantitative synthesis was structured around four main outcomes: preterm birth, low birth weight, SGA, and cognitive/global development in childhood. The first three outcomes were analyzed as binary variables on the logRR scale, which was the primary analytical measure at all stages of modeling and presentation. Cognitive/global development was analyzed as a continuous variable and harmonized using the standardized mean difference with Hedges correction (g). All analyses used random-effects models, with between-study variance estimated by REML and pooled-effect inference performed using KH adjustment.

The analyses included nine estimates for preterm birth, nine for low birth weight, six for SGA, and five for cognitive/global development. Some cohorts contributed to more than one perinatal outcome because preterm birth, birth weight, and fetal growth adequacy were evaluated in the same participants. However, each meta-analysis was conducted separately using only estimates corresponding to the specific outcome.

The direction of effects was harmonized a priori. For binary outcomes, a logRR greater than 0 indicated a higher frequency of the adverse event among mothers with deficiency or lower vitamin B12 availability, whereas logRR equal to 0 represented the null value. For cognitive/global development, positive Hedges g values were oriented to indicate better performance among children exposed to higher maternal vitamin B12 availability or supplementation. This standardization kept the direction of individual and pooled estimates consistent across studies.

Table 1 summarizes the results of the four main meta-analyses.

**Table 1.** Summary of meta-analyses of maternal vitamin B12 deficiency or exposure and perinatal and neurodevelopmental outcomes.

| Outcome | k | Pooled effect | 95% CI (KH) | p | Heterogeneity |
| --- | --- | --- | --- | --- | --- |
| Preterm birth | 9 | logRR = 0.190 | -0.046 to 0.427 | 0.101 | tau2 = 0.013; I2 = 19.2%; Q = 9.91 |
| Low birth weight | 9 | logRR = 0.136 | -0.013 to 0.286 | 0.069 | tau2 = 0.000; I2 = 4.8%; Q = 8.40 |
| SGA | 6 | logRR = 0.047 | -0.083 to 0.177 | 0.397 | tau2 = 0.000; I2 = 0%; Q = 2.97 |
| Cognitive/global development | 5 | Hedges g = 0.266 | 0.034 to 0.499 | 0.034 | tau2 = 0.020; I2 = 55.7%; Q = 9.03 |

Source: prepared by the authors based on the included studies.
Note: for the three binary outcomes, the pooled effect is presented as logRR; for cognitive/global development, it is presented as Hedges g. The null value is 0 on both scales.

### 3.3 Preterm birth

Nine estimates were combined in the analysis of preterm birth. This group included the cohorts by Baker, Bergen, Bhate, Chen, Dwarkanath, Kaymaz, Krishnaveni, Sukumar, and Yajnik. Individual estimates showed variable magnitudes, although most were concentrated around values close to or slightly above the null (Rogne *et al*., 2017).

Individual logRRs ranged from –0.580 to 1.723. Among the more precise estimates, Bergen showed a logRR of 0.010 (95% CI: –0.236 to 0.255), Chen 0.525 (95% CI: 0.020 to 1.026), Krishnaveni 0.351 (95% CI: –0.117 to 0.815), and Yajnik 0.270 (95% CI: –0.139 to 0.678). Sukumar showed the highest point estimate, logRR 1.723, but with a wide 95% CI (–0.342 to 3.781), resulting in lower precision in the model.

The pooled effect under the random-effects model was logRR 0.190, with a KH 95% CI from –0.046 to 0.427 and p = 0.101. Because the logRR scale has a null value of 0, the interval included no association. For complementary clinical interpretation, exponentiation of the point estimate corresponds approximately to 1.21, without changing the fact that the primary analysis and inference were performed on the logRR scale.

Heterogeneity was low. Between-study variance estimated by REML was tau2 = 0.013, while I2 was 19.2%. Cochran Q was 9.91, without statistical evidence of heterogeneity (p = 0.272). The distribution of individual estimates and the pooled effect is shown in Figure A1, available in Appendix A.

### 3.4 Low birth weight

The low birth weight meta-analysis also included nine estimates from the same main cohorts used in the preterm birth analysis. Individual logRRs ranged from –0.693 to 1.619, although estimates with greater statistical weight were concentrated closer to 0 (Rogne *et al*., 2017).

Bergen showed a logRR of 0.095 (95% CI: –0.139 to 0.336), Bhate 0.131 (95% CI: –0.400 to 0.658), Chen 0.519 (95% CI: –0.020 to 1.051), Dwarkanath 0.182 (95% CI: –0.186 to 0.554), Krishnaveni 0.199 (95% CI: –0.105 to 0.513), and Yajnik 0.068 (95% CI: –0.151 to 0.293). Kaymaz had the largest point estimate, logRR 1.619, but with a wide 95% CI (–0.261 to 3.497), indicating high imprecision.

The pooled effect was logRR 0.136, with a KH 95% CI from –0.013 to 0.286 and p = 0.069. The estimate remained in the direction of greater occurrence of low birth weight in the group with lower maternal vitamin B12 availability, but the 95% CI included 0. Exponentiation of the point estimate, presented only as complementary interpretation, corresponds approximately to 1.15.

Heterogeneity was minimal. REML estimation resulted in tau2 = 0.000, with I2 = 4.8%. The Q statistic was 8.40 (p = 0.395), showing high consistency among individual estimates. The complete forest plot for this outcome is presented in Figure B1, Appendix B.

### 3.5 Small for gestational age

Six studies provided quantitatively comparable estimates for SGA. The cohorts by Baker, Bergen, Chen, Dwarkanath, Krishnaveni, and Sukumar were included. This outcome showed a particularly homogeneous distribution, with most estimates concentrated around the null value (Rogne *et al*., 2017).

Individual logRRs were –0.198 for Baker, –0.030 for Bergen, 0.030 for Chen, 0.278 for Dwarkanath, 0.068 for Krishnaveni, and 0.113 for Sukumar. The corresponding 95% CIs overlapped widely and, in all cases, included 0.

The REML model yielded a pooled logRR of 0.047, with a KH 95% CI from –0.083 to 0.177 and p = 0.397. Thus, the pooled estimate remained close to the null value on the analytical scale itself. The exponentiated point estimate corresponds approximately to 1.05 and is presented only as complementary interpretive information.

No heterogeneity was identified across studies. Between-study variance was tau2 = 0.000 and I2 = 0%. Cochran Q was 2.97 (p = 0.704). The graphical representation of the six individual estimates and the pooled effect is shown in Figure C1, available in Appendix C.

### 3.6 Cognitive and global neurodevelopment

Five independent studies contributed to the outcome of cognitive/global development: Bhate, Chandyo, MATCOBIND, Lai, and PRIYA/D’Souza. Unlike the perinatal outcomes, these studies used different instruments and developmental scales. To enable pooling, results were converted to Hedges g, with positive values indicating better performance associated with greater maternal exposure to vitamin B12 (Bhate *et al*., 2012; Chandyo *et al*., 2023; D’Souza *et al*., 2021; Lai *et al*., 2019; Nagpal *et al*., 2026).

Individual estimates ranged from approximately 0.06 to 0.59 standard deviations. Bhate showed an effect of approximately g = 0.39, whereas Chandyo showed an estimate close to the null, g = 0.06. In MATCOBIND, the standardized difference was approximately 0.24; Lai showed an effect of approximately 0.42; and PRIYA/D’Souza showed the largest estimate, about 0.59, although with greater uncertainty.

The pooled effect was Hedges g = 0.266, with a KH 95% CI from 0.034 to 0.499 and p = 0.034. The pooled estimate therefore corresponded to a small positive standardized difference in cognitive/global development associated with higher maternal vitamin B12 availability or supplementation.

Heterogeneity was moderate. Between-study variance was tau2 = 0.020, while I2 was 55.7%. The Q statistic was 9.03, with p = 0.060. The distribution of individual effects shows variation in magnitude across populations and instruments, but predominance of estimates in the positive direction. The corresponding forest plot is provided in Figure D1, Appendix D.

### 3.7 Secondary outcomes

Head circumference, motor development, language, and other neonatal parameters remained secondary outcomes. These outcomes were reported heterogeneously across studies, including absolute means, regression coefficients, correlations, standardized results, and assessments at different ages. For this reason, they were not incorporated into the four main quantitative groupings (Cruz-Rodriguez *et al*., 2023; Srinivasan *et al*., 2017; Thomas *et al*., 2019).

Head circumference was evaluated in different cohorts, but the way results were reported did not allow harmonization with the same level of comparability obtained for preterm birth, low birth weight, and SGA. Similarly, motor and language outcomes appeared in some neurodevelopmental studies, but with fewer independent estimates and distinct instruments.

Thus, the main analysis focused on the four outcomes with the greatest availability and quantitative comparability. Overall, results showed positive point estimates for preterm birth and low birth weight, absence of a relevant association for SGA, and a small positive standardized effect for cognitive/global development. Heterogeneity varied substantially across domains, being practically absent in anthropometric outcomes and higher in the neurodevelopmental grouping.

## 4 DISCUSSION

This meta-analysis gathered evidence on the relationship between maternal vitamin B12 status and perinatal and neurodevelopmental outcomes, organizing the quantitative synthesis into four main domains: preterm birth, low birth weight, SGA, and cognitive/global development. For the three binary outcomes, statistical interpretation was conducted primarily on the logRR scale, whose null value is 0. Results showed distinct patterns: preterm birth and low birth weight had positive pooled estimates, but their KH-adjusted 95% CIs crossed 0; SGA had an estimate close to the null; and cognitive/global development showed a small positive standardized effect associated with higher maternal vitamin B12 availability or supplementation.

In the preterm birth analysis, the pooled logRR was 0.190 (KH 95% CI: –0.046 to 0.427; p = 0.101). The point estimate was in the direction of a higher occurrence of preterm birth among mothers with lower B12 availability, but statistical uncertainty precludes interpretation as a conclusive association. Low heterogeneity, with I2 = 19.2%, indicates that the direction and magnitude of estimates were relatively consistent across cohorts. Exponentiation of the point effect corresponds approximately to 1.21 and may help clinical interpretation, but it does not replace logRR as the primary measure of this meta-analysis (Rogne *et al*., 2017).

Vitamin B12 participates in one-carbon metabolism, DNA synthesis, and remethylation of homocysteine to methionine. Alterations in these pathways may affect cell proliferation, placental metabolism, and availability of methyl groups required for epigenetic regulation. Therefore, there is biological plausibility for an association between maternal B12 deficiency and obstetric outcomes. However, maternal B12 status is also related to dietary patterns, prenatal supplementation, socioeconomic conditions, folate status, body mass index, and other factors that may influence preterm birth risk. For this reason, the pooled estimate should be understood as an average association obtained across distinct clinical and epidemiological contexts, rather than as the isolated effect of the micronutrient (Green *et al*., 2017; Pepper and Black, 2011).

For low birth weight, the pooled logRR was 0.136 (KH 95% CI: –0.013 to 0.286; p = 0.069). The result remained close to the null threshold but showed minimal heterogeneity, with I2 = 4.8% and tau2 estimated at zero. This pattern suggests high agreement across studies regarding effect magnitude on the logarithmic scale, even though the 95% CI included 0. Low birth weight is a particularly complex outcome because it may result from either preterm birth or fetal growth restriction. Thus, even if B12 deficiency is related to one of these mechanisms, combining both into a single low-birth-weight definition may dilute specific associations (Rogne *et al*., 2017).

This interpretation is reinforced by the result observed for SGA. In this analysis, the pooled logRR was 0.047 (KH 95% CI: –0.083 to 0.177), accompanied by I2 equal to zero. The proximity of the estimate to 0 and the absence of heterogeneity suggest that, within the analyzed populations, there was no consistent evidence of an association between lower maternal B12 availability and inadequate fetal growth relative to gestational age. Therefore, the slight elevation observed for low birth weight may not be predominantly explained by intrauterine growth restriction. Differences in gestational duration may contribute more importantly to this result, a hypothesis coherent with the direction observed in the preterm birth analysis (Rogne *et al*., 2017).

Cognitive/global development behaved differently from the perinatal outcomes. The pooled effect was Hedges g = 0.266, with a 95% CI from 0.034 to 0.499 and p = 0.034, indicating a small standardized difference favoring higher maternal availability or supplementation of vitamin B12. Although the magnitude was modest, this finding is relevant because the studies used distinct assessment instruments and followed children at different ages. Moderate heterogeneity, with I2 = 55.7%, is compatible with this methodological diversity (Bhate *et al*., 2012; Chandyo *et al*., 2023; D’Souza *et al*., 2021; Lai *et al*., 2019; Nagpal *et al*., 2026).

The association with neurodevelopment has consistent biological plausibility. Vitamin B12 is required for methionine synthesis and maintenance of myelin, and it participates in processes related to cell division and nervous system maturation. During pregnancy and the first years of life, periods characterized by rapid structural and functional brain organization, inadequate availability of this micronutrient may theoretically interfere with myelination, neuronal growth, and circuit formation. These mechanisms make it biologically plausible that differences in maternal B12 status may be reflected in child developmental measures, even when some anthropometric outcomes at birth remain close to the null (Green *et al*., 2017; Pepper and Black, 2011).

At the same time, the cognitive result should be interpreted considering differences between observational exposure and experimental supplementation. Some studies assessed maternal B12 concentrations, whereas others investigated interventions delivered before or during pregnancy. These designs address related but not identical questions. Low maternal concentrations may reflect dietary intake, absorption, body stores, and metabolic conditions; supplementation trials evaluate deliberate modification of this exposure. Combining them on a standardized scale allows assessment of the overall direction of the phenomenon, but does not eliminate this etiological heterogeneity (Bonifacio *et al*., 2025; Chandyo *et al*., 2023; D’Souza *et al*., 2021; Nagpal *et al*., 2026).

Another relevant aspect is the age at which neurodevelopment was evaluated. Studies conducted in early infancy use scales oriented toward initial abilities, whereas later assessments incorporate language, memory, attention, and more complex cognitive functions. As the interval between gestational exposure and child assessment increases, the influence of postnatal factors also increases, including feeding, schooling, family stimulation, socioeconomic status, and intercurrent clinical conditions. These factors probably contribute to the greater heterogeneity observed for the cognitive outcome compared with the perinatal outcomes (Cruz-Rodriguez *et al*., 2023; D’Souza *et al*., 2021; Lai *et al*., 2019).

From a methodological perspective, the use of random-effects models with REML estimation and KH adjustment allowed uncertainty between studies to be incorporated more conservatively. Choosing logRR as the primary measure for binary outcomes also preserved a scale symmetric around the null value of 0 during estimation. This choice was particularly important for preterm birth and low birth weight, where point estimates were positive but adjusted intervals included 0. Interpretation should therefore consider logRR magnitude, 95% CI, heterogeneity, and consistency across studies simultaneously, rather than relying only on the p value (Hartung and Knapp, 2001; Viechtbauer, 2005).

Limitations of this synthesis include differences in definitions of vitamin B12 deficiency, variation in timing of maternal collection, population differences, and the combination of observational studies and interventions. In addition, some secondary outcomes, such as head circumference, language, and motor development, were presented in heterogeneous statistical formats and could not be incorporated with the same level of comparability into the main models. This information remains relevant for the global characterization of evidence, but requires narrative interpretation or specific analyses (Bonifacio *et al*., 2025; Rogne *et al*., 2017).

Taken together, the results suggest a pattern in which maternal vitamin B12 deficiency or lower availability shows a modest association with preterm birth and low birth weight, no consistent evidence of a relationship with SGA, and a small beneficial association between greater maternal availability or supplementation and cognitive/global development in childhood. Low heterogeneity in perinatal outcomes contrasts with moderate variability in neurodevelopment, indicating that the effects of vitamin B12 may depend on the outcome analyzed, the exposure period, and the timing of child assessment (Bonifacio *et al*., 2025; Rogne *et al*., 2017).

## 5 CONCLUSION

In this systematic review and meta-analysis, lower maternal vitamin B12 availability showed positive pooled logRRs for preterm birth and low birth weight, although adjusted 95% CIs included 0. For SGA, the pooled logRR remained close to the null. For cognitive/global development, higher maternal vitamin B12 availability or supplementation was associated with a small favorable standardized effect, with moderate heterogeneity.

The findings indicate that the effects of vitamin B12 may vary according to the outcome, exposure timing, and clinical and nutritional characteristics of the studied populations. Future studies using standardized definitions of deficiency, longitudinal measurement of maternal status, and harmonized neurodevelopmental assessment may help clarify the magnitude and clinical relevance of these associations.

## DECLARATIONS

### Ethics approval

Not applicable. This study used aggregate data from previously published studies.

### Data availability

All data used and analyzed in this review and meta-analysis are available within the manuscript and its appendices.

### Author contributions – Contributor Roles Taxonomy (CRediT)

**Eduarda Rodrigues Belfort de Carvalho:** Conceptualization, Methodology, Investigation, Data curation, Formal analysis, Visualization, Writing – original draft, Writing – review & editing.

**Hércules Kanaan Pereira Sousa:** Conceptualization, Methodology, Formal analysis, Software, Visualization, Data curation, Writing – original draft, Writing – review & editing, Project administration.

**Thiago de Sousa Siqueira:** Methodology, Investigation, Data curation, Validation, Writing – original draft, Writing – review & editing.

**Ricardo Rezende de Queiroz:** Investigation, Data curation, Validation, Writing – review & editing.

**Juliana Borges Tavares da Silva:** Investigation, Data curation, Validation, Writing – review & editing.

**Ana Júlia Hortegal Cavalcante:** Investigation, Data curation, Validation, Writing – review & editing.

**Paloma Layonara Vieira Viana:** Investigation, Validation, Writing – review & editing.

**Maria Eduarda Araújo Magalhães:** Investigation, Validation, Writing – review & editing.

**Gabriel Segrini Fidencio:** Validation, Writing – review & editing.

**Ana Raquel Batista de Carvalho:** Supervision, Conceptualization, Methodology, Project administration, Validation, Writing – review & editing.

### Author consent for preprint

All authors have approved this version of the manuscript and consent to its submission as a medRxiv preprint

## APPENDIX A – PRETERM BIRTH

**Figure A1.**
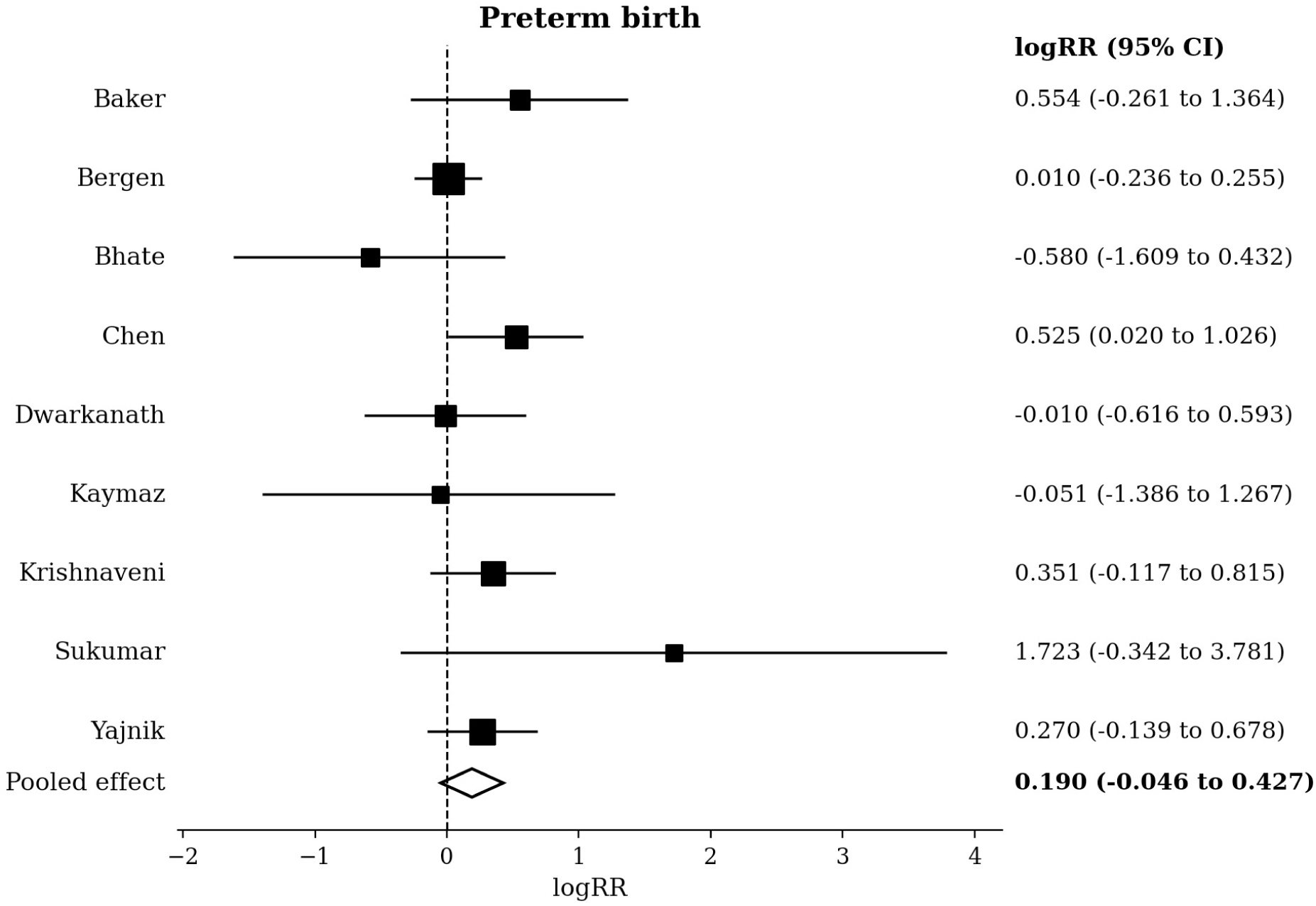
Forest plot of logRR estimates for preterm birth. Source: prepared by the authors based on the included studies. Note: random-effects model; pooled effect logRR = 0.190 (95% CI KH: –0.046 to 0.427).

## APPENDIX B – LOW BIRTH WEIGHT

**Figure B1.**
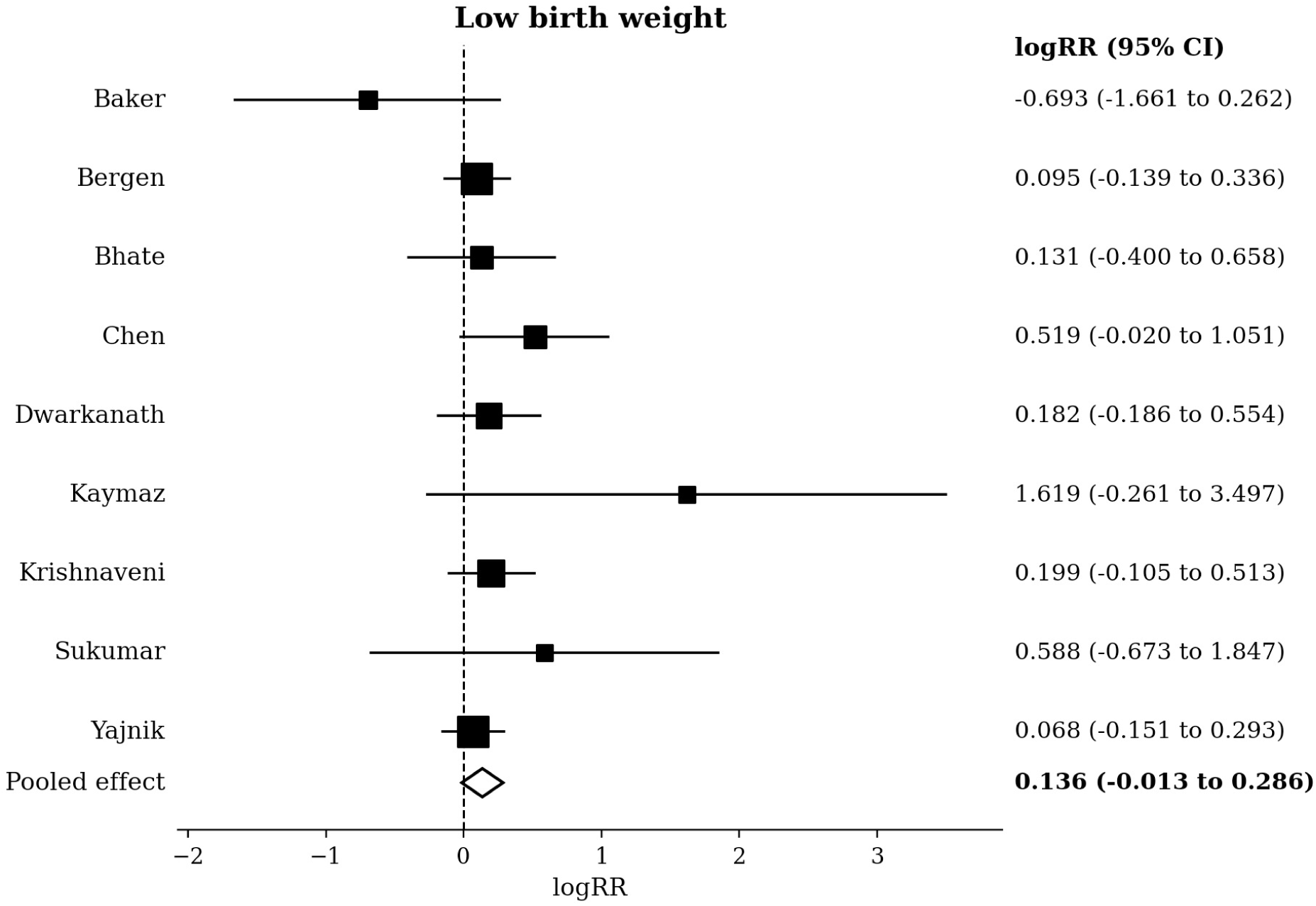
Forest plot of logRR estimates for low birth weight. Source: prepared by the authors based on the included studies. Note: random-effects model; pooled effect logRR = 0.136 (95% CI KH: –0.013 to 0.286).

## APPENDIX C – SMALL FOR GESTATIONAL AGE

**Figure C1.**
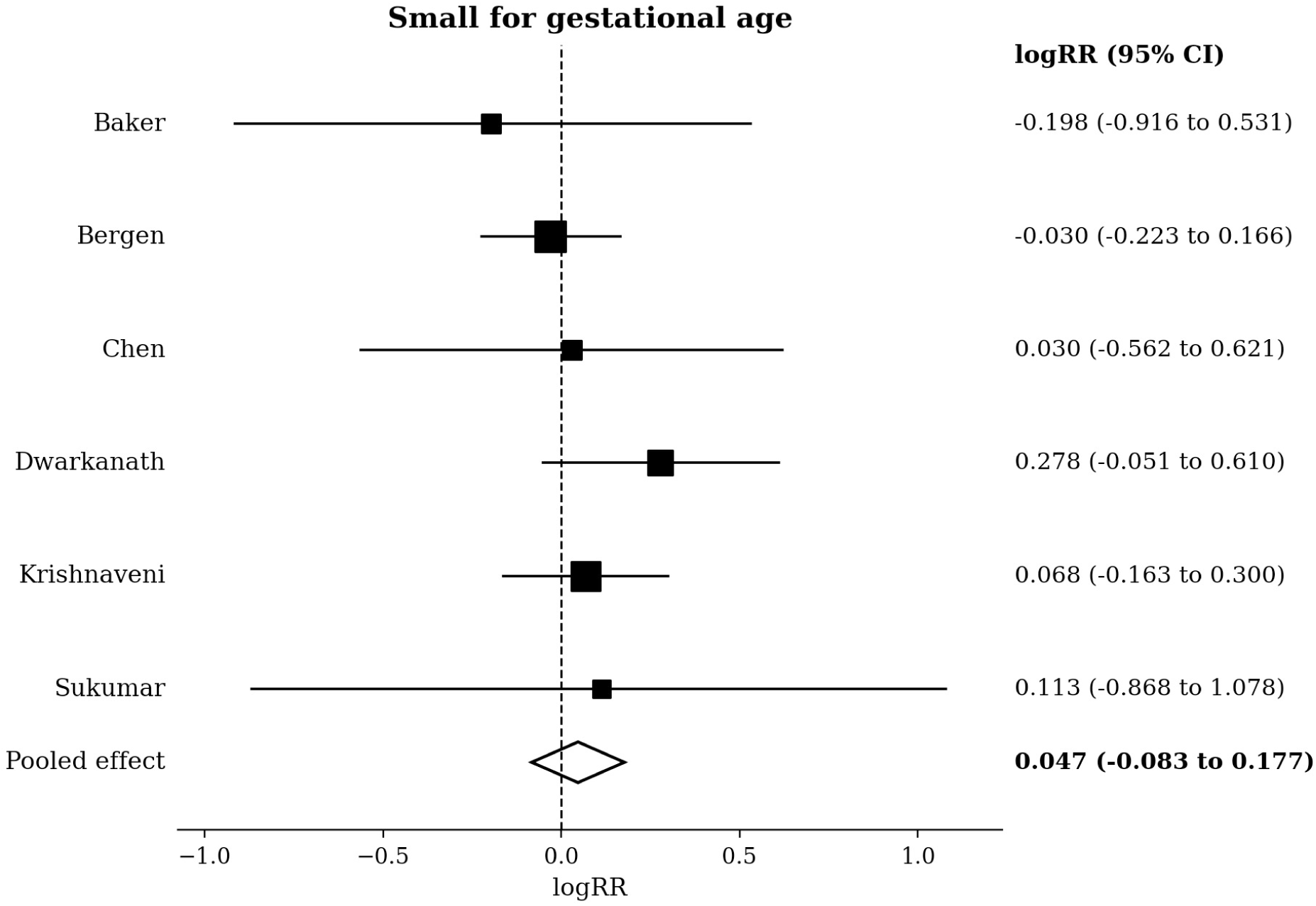
Forest plot of logRR estimates for SGA. Source: prepared by the authors based on the included studies. Note: random-effects model; pooled effect logRR = 0.047 (95% CI KH: –0.083 to 0.177).

## APPENDIX D – COGNITIVE/GLOBAL DEVELOPMENT

**Figure D1.**
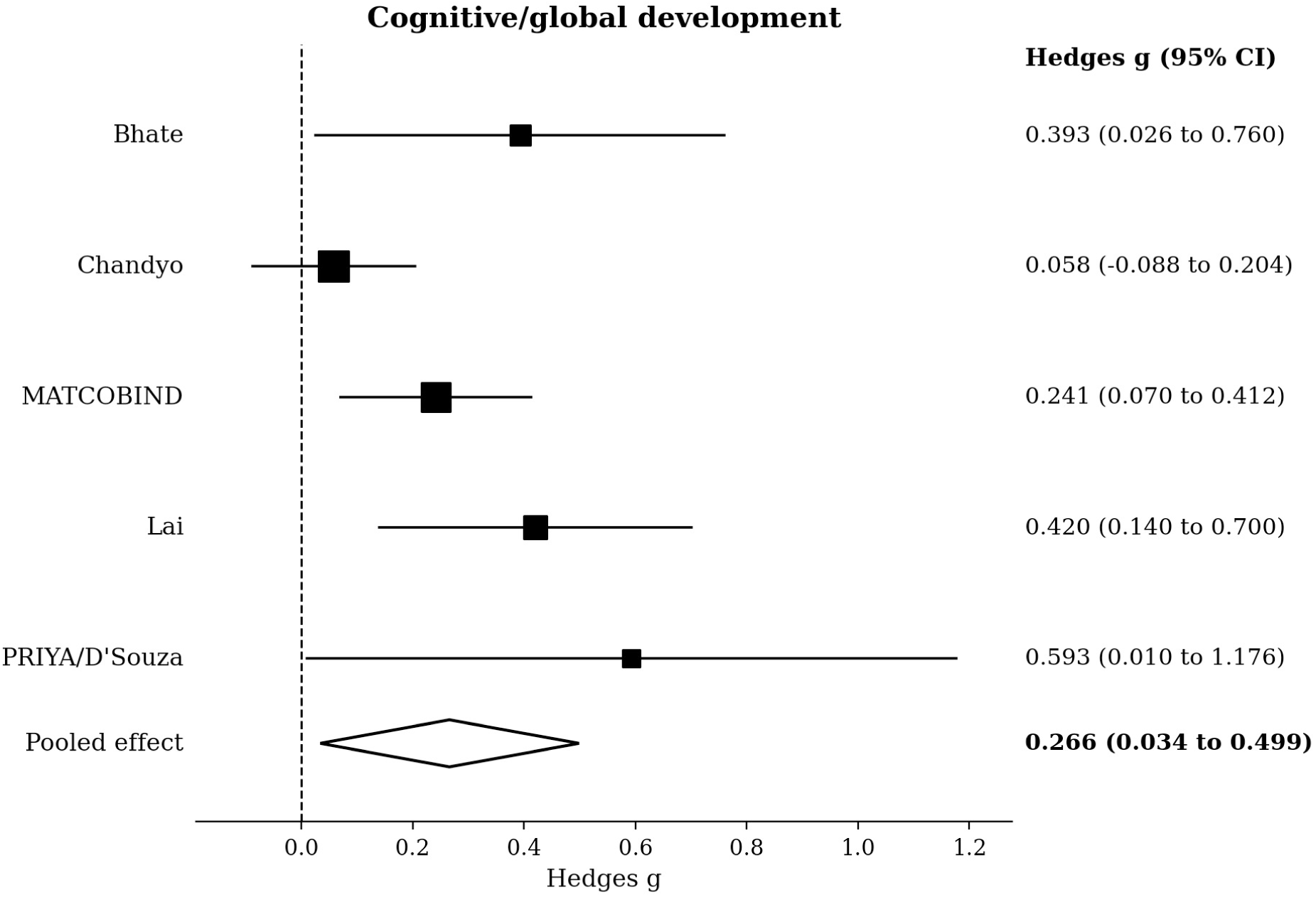
Forest plot for cognitive/global development on the Hedges g scale. Source: prepared by the authors based on the included studies. Note: random-effects model; pooled effect Hedges g = 0.266 (95% CI KH: 0.034 to 0.499). Positive values favor greater maternal B12 availability or supplementation.

## APPENDIX E – PRISMA 2020 FLOW DIAGRAM

**Figure E1.**
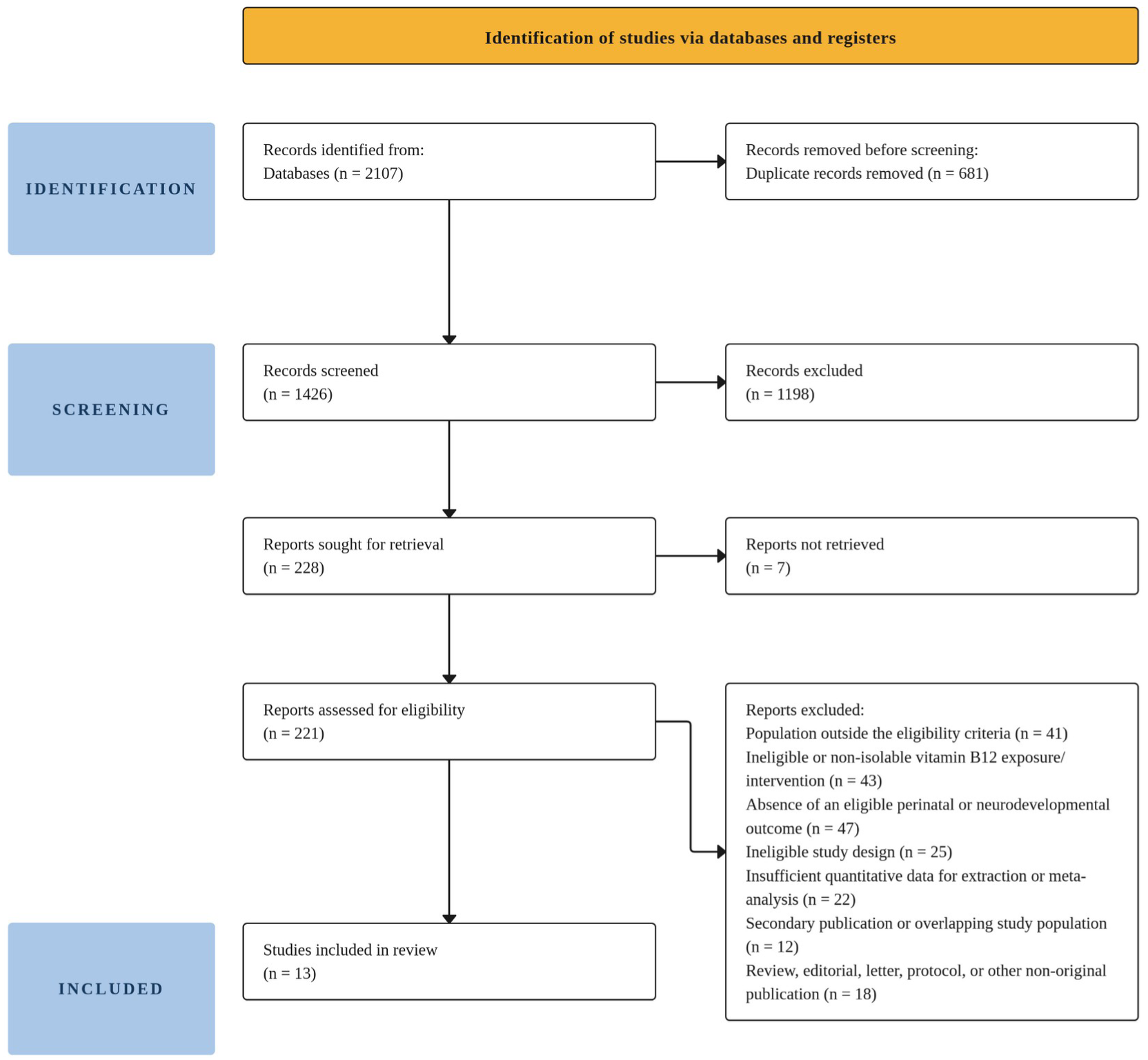
PRISMA 2020 flow diagram of study identification, screening, eligibility assessment, and inclusion. Source: prepared by the authors according to PRISMA 2020.

